# Associations of habitual fish oil use with risk of SARS-CoV-2 infection and COVID-19-related outcomes in UK: national population based cohort study

**DOI:** 10.1101/2022.09.14.22279933

**Authors:** Yuying Ma, Lijun Zhang, Ruijie Zeng, Dongling Luo, Rui Jiang, Huihuan Wu, Zewei Zhuo, Qi Yang, Jingwei Li, Felix W Leung, Jinghua Wang, Weihong Sha, Hao Chen

## Abstract

**Objectives:** To prospectively investigate the associations of habitual fish oil use with Severe acute respiratory syndrome coronavirus 2 (SARS-CoV-2) infection, hospital admission, or mortality with Corona Virus Disease-19 (COVID-19) in a large-scale cohort.

**Design:** Prospective population-based cohort study.

**Setting:** UK Biobank.

**Participants:** A total of 110 440 participants aged 37 -73 years who completed a questionnaire on supplement use, which included fish oil at baseline were enrolled between 2006 and 2010 and followed up until 2022.

**Main exposure:** All participants filled out questionnaires about the habitual use of supplements, including fish oil.

**Main outcome measures:** SARS-CoV-2 infection, COVID-19 hospital admission and COVID-19 mortality.

**Results:** At baseline, 29 424 (26.6%) of the 110 440 participants reported habitual use of fish oil supplements. The multivariable adjusted hazard ratios for habitual users of fish oil versus non-users were 0.95 (0.93 to 0.98) for SARS-CoV-2 infection among participants with follow-up time less than 12.1 years but no significant associations were observed for participants with follow-up time more than 12.1 years. For COVID-19-related outcomes, the hazard ratios were 0.79 (95% confidence interval 0.71 to 0.88) for COVID-19 hospital admission and 0.72 (0.60 to 0.87) for COVID-19 mortality. For COVID-19-related outcomes, the association seemed to be stronger among those with longstanding illness. The Cox proportional hazard analysis after propensity-score matching yielded consistent results.

**Conclusions:** Habitual fish oil supplement is associated with a lower risk of hospital admission and mortality with COVID-19, but not associated with SARS-CoV-2 infection in the population with more than 12.1 years of follow-up.

## 1. Introduction

Fish oil has been recommended for daily use because of its important role in preventing cardiovascular disease^1^. Accordingly, fish oil supplement has been widely used for decades, particularly in developed countries^2^. It has been estimated that approximately 22% of adults in New Zealand use fish oil^3^.

Coronavirus disease 2019 (COVID-19) is caused by the newly identified Severe acute respiratory syndrome coronavirus 2 (SARS-CoV-2)^4^. Since it first broke out on December 31, 2019, COVID-19 has spread around the world at outrageous speed^5^. As of 19 August 2022, there have been 591.7 million confirmed cases of COVID-19 globally, including 6.4 million deaths^6^. Notably, the emergence of multiple variants of SARS-CoV-2, varying in pathogenic features, threatens public health unprecedentedly^7^. Considering the devastating impact of the COVID-19 pandemic, there is still a further concern for developing specific treatments to minimize the risk of COVID-19.

Mounting evidence supports a beneficial role of fish oil intake in regulating immune function, which helps reduce the risk of SARS-CoV-2-infection and increase survival rate in COVID-19 patients^8 9^. As such, fish oil is thought to be feasible to prevent and treat COVID-19^10 11^. However, most recent reports are randomized controlled trials with inadequate sample size, making it difficult to apply their findings to larger populations^12^. In this regard, large-scale population-based cohort studies in real-life setting are still warranted to evaluate the effectiveness of fish oil supplement.

In this large-scale prospective cohort study, we assessed the association between habitual fish oil use and the risks of SARS-CoV-2 infection, hospital admission and mortality with COVID-19, taking advantage of UK Biobank with half a million UK adults involved and providing updated data on COVID-19 events.

## 2. Methods

### 2.1 Study population and design

The UK Biobank is a large prospective cohort study that recruited 502 413 people aged 37 to 73 years between 2006 and 2010 in the UK (https://www.ukbiobank.ac.uk/)^13^. Written informed consent was acquired from each participant, and ethical approval was obtained from the North West Multi-Center Research Ethics Committee (approval number: 11/NW/0382, 16/NW/0274, and 21/NW/0157). The current study has been approved under the UK Biobank project 83339. Participants were recruited from one of 22 assessment centers across England, Scotland, and Wales, where they completed a detailed touch screen questionnaire, provided biological samples, and took a set of physical measurements at baseline. a series of physical measurements were also taken. All participants were required to provide written informed consent. Inpatient hospital data were mapped across England, Scotland and Wales based on Hospital Episode Statistics (HES) in England, the Scottish Morbidity Record, and the Patient Episode Database for Wales. Data on mortality are updated from the National Health Service (NHS) Digital for participants in England and Wales and from the NHS Central Register for participants in Scotland. The UK Biobank has also been linked to Public Health England (PHE), Public Health Scotland (PHS) and SAIL after the pandemic of COVID-19 for getting results of COVID-19 tests in English clinical diagnostics laboratories (based on RT-PCR of nose/throat swab samples and – the Thriva coronavirus antibody test, available since 16st March) for the purpose of facilitating COVID-19-related research^14^. The diagnoses in inpatient hospital data in England have been widely validated in previous studies, showing consistently high quality and diagnostic accuracy. Participants with incomplete data on the use of fish oil (n=6 187), those with incomplete COVID-19 testing data (n=300 502), those who subsequently withdrew from the study (n=1 298) and with incomplete covariate data (n=83 986) were excluded from the analysis. In total, our analysis included 110 440 participants **(Figure 1**). In addition, for COVID-19 hospital admission, we excluded participants who died from the virus without being hospitalized.

**Figure 1.**
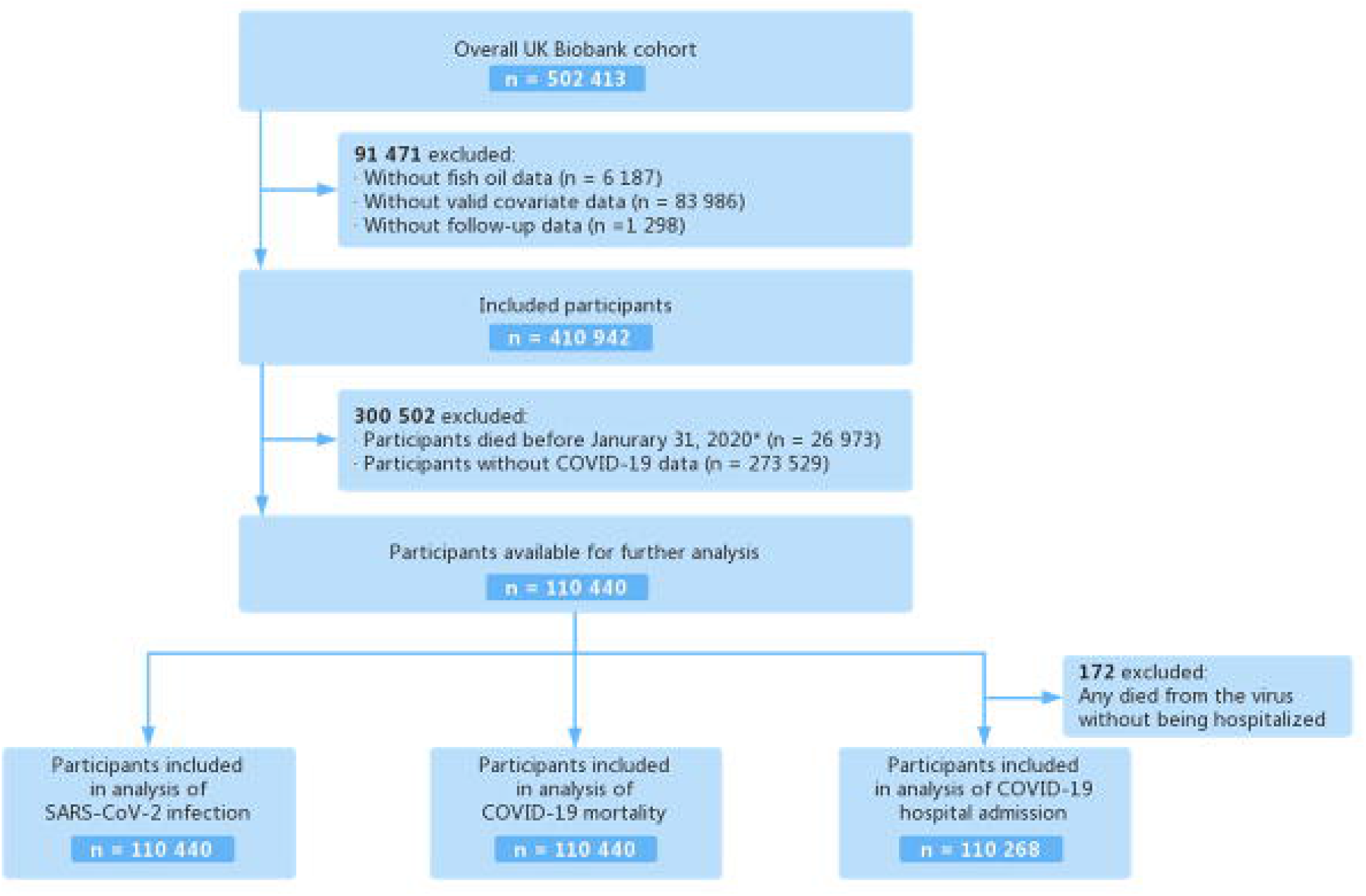
Flow chart of eligible participants selection. *The first COVID-19 case was diagnosed on Jan 31, 2020, in the UK.

### 2.2 Assessment of fish oil supplement use

The habitual use of fish oil supplement was recorded using a touchscreen questionnaire at baseline, which has been shown good agreement and repeatability to the later 24-hour dietary assessment. Participants were asked, “Do you regularly take any of the following? (You can select more than one answer)”. We scored habitual use of fish oil supplements as “1=yes” or “0=no”^15^.

### 2.3 Assessment of COVID-19

SARS-CoV-2 infection was considered by self-antibody tests reports of blood samples and nose/throat swab tests reports. In addition, participants died or were hospitalized due to COVID-19 were also defined as COVID-19 infected even they have no COVID-19 test reports.

We defined COVID-19 hospital admission and mortality as an inpatient or death with COVID-19 (ICD-10 U07.1 and U07.2) as the cause according to data from the death registers and hospital inpatient (updated up to September 30, 2021).

Follow-up duration started at the date when participants attended the baseline survey at the assessment center and ended at the time of COVID-19 diagnosis, lost to follow-up, inpatient or mortality whichever occurred first.

### 2.4 Covariates

Baseline questionnaire were used to assess several possible confounding variables: sociodemographic factors (age, sex, ethnicity, and household income), socioeconomic status (Townsend Deprivation Index), lifestyle habits (smoking status, alcohol consumption), body mass index (BMI), physical activity, dietary intake (raw vegetables, fresh fruit, oily fish and non-oily fish), comorbidities (hypertension, type-2 diabetes, myocardial infraction, renal failure, stoke, asthma, chronic obstructive pulmonary disease, and longstanding illness), vitamin supplementation (vitamin A, vitamin B, vitamin C, vitamin D, vitamin E, multivitamin, or folic acid), and mineral and other dietary supplementation (calcium, iron, zinc, or selenium). The Townsend Deprivation Index, used as an indicator of socioeconomic status, is derived from the residential postcode and is provided directly from the UK Biobank. Information on lifestyle and dietary intake is collected by touchscreen questionnaire at baseline. Physical activity is calculated by Metabolic Equivalent Task (MET) minutes per week for moderate activity. Data on comorbidities are self-reported by participants in Verbal interview. Details of these assessments can be found on the UK Biobank website (www.ukbiobank.ac.uk).

### 2.5 Statistical analyses

The baseline characteristics were showed by percentages for categorical variables and mean (standard deviation [SD]) for continuous variables according to the distribution of data. The associations between habitual fish oil use and outcomes (SARS-CoV-2 infection, COVID-19 hospital admission and mortality) were explored using Cox proportional hazard models. The proportional hazard assumption was evaluated by tests based on Schoenfeld residuals and no evidence of violation was observed except SARS-CoV-2 infection. To comply with proportional hazard assumption, time at risk of SARS-CoV-2 infection was split into <12.1 years and ≥12.1 years, which is the median follow-up time and also the time of outbreak of the new coronavirus strain, Omicron in UK. Three sets of models were used. The basic model (model l) was non-adjusted. The preliminary adjusted model (model 2) for baseline age (years) and sex (male or female). The multivariable model (model 3) was adjusted for additional variables, including the Townsend Deprivation Index, ethnicity (white, other race, Asian, black, Indian, Pakistani, Bangladeshi, or any other Asian background except Chinese, mixed, or other ethnic group), household income (<£18 000, £18 000-£30 999, £31 000-£51 999, £52 000-£100 000, or >£100 000), BMI, smoking status (never, previous or current), alcohol consumption (never, special occasions, one to three times a month, once or twice a week, three or four times a week, daily or most daily), physical activity (MET minutes/week), fresh fruit consumption (pieces/day), raw vegetable consumption (tablespoons/day), oily fish consumption (<2 or ≥2 times/week), non-oily fish consumption (<2 or ≥2 times/week), vitamin supplementation (yes or no), mineral and other dietary supplementation (yes or no), hypertension (yes or no), type 2 diabetes (yes or no), myocardial infraction (yes or no), renal failure(yes or no),stroke(yes or no), COPD(yes or no), asthma(yes or no), longstanding illness (yes or no).

Furthermore, inverse probability of treatment weighting that was based on propensity score was used to construct weighted cohorts of patients who differed with fish oil use but were similar with respect to other measured characteristics. And weighted Kaplan-Meier curves were constructed to compare the event -free probability of habitual fish oil users and non-users.

As an additional approach to adjust covariates, matching on propensity scores for fish oil exposure was carried out. We matched fish oil users to non-users individuals at a 1:1 ratio using a greedy nearest neighbor method with the MATCHIT package in R^16^. The overall quality of the matched sample was assessed by comparing the standardized mean differences of all covariates and by visual inspection of propensity score distributions between unmatched and matched samples.

We performed a stratified analysis to estimate potential modification effects according to sex (male or female), age (<60 or ≥60 years), BMI (≥30 or <30), alcohol consumption(never or special occasions only or ≥1 time/month), smoking status(never, previous or current), fruit consumption(<mean or ≥mean), oily fish consumption (<2 or ≥2 times/week), non-oily fish consumption (<2 or ≥2 times/week), mineral and other dietary supplementation(yes or no), hypertension (yes or no), myocardial infraction (yes or no), stroke (yes or no), asthma (yes or no), longstanding illness(yes or no). All analyses were performed using R software (version 4.2.1, https://www.r-project.org/) in Rstudio. We considered a P value less than 0.05 (two-sided) to be statistically significant.

## 3. Results

### 3.1 Baseline characteristics

The baseline characteristics of the participants showed in **Table 1** are stratified by fish oil use status (users versus nonusers). Of the 110 440 UK biobank participants,60 026 (54.44%) were female and the mean age is 55.5 years. Overall, 29 424(26.6%) of the participants reported the use of fish oil supplementation at baseline. Fish oil users in the study were older and more female, previous smokers, had a higher household income, and took more physical activity compared with nonusers. In addition, they were also more likely to take more fresh fruit, raw vegetables and oily fish and habitually took vitamin and mineral supplements. Moreover, fish oil users were observed to have a higher prevalence of hypertension, myocardial infraction and longstanding illness, but a lower prevalence of asthma.

**Table 1.** Baseline characteristics of UK Biobank participants by fish oil use.

| Characteristics | Fish oil user | Fish oil non-user | Overall |
| --- | --- | --- | --- |
| **Number of participants, n(%)** | 29 424(26.6) | 81 016(73.4) | 110 440(100.0) |
| **Age, mean(SD), years** | 58.1(7.02) | 54.6(7.74) | 55.5(7.71) |
| **Sex, female, n(%)** | 16589(56.4) | 43 437(53.6) | 60 026(54.4) |
| **Ethnicity, White, n(%)** | 28710(97.6) | 78 634(97.1) | 107 344(97.2) |
| **Household income (£\*)** |  |  |  |
| <18 000, n(%) | 4 389(14.9) | 9 598(11.8) | 13 987(12.6) |
| 18 000-30 999, n(%) | 7 061(24.0) | 15 411(19.0) | 22 472(20.3) |
| 31 000-51 999, n(%) | 7 479(25.4) | 21 520(26.6) | 28 999(26.3) |
| 52 000-100 000, n(%) | 5 902(20.1) | 21 031(26.0) | 26 933(24.4) |
| >100 000, n(%) | 1 478(5.0) | 6 558(8.1) | 8 036(7.3) |
| Missing, n(%) | 3 115(10.6) | 6 898(8.5) | 10 013(9.1) |
| **Deprivation index, mean(SD)** | -1.93(2.70) | -1.69(2.9) | -1.76(2.8) |
| **BMI, mean(SD), kg/m<sup>2</sup>** | 26.58(4.3) | 26.95(4.7) | 26.85(4.6) |
| **Alcohol consumption** |  |  |  |
| Daily or almost daily, n(%) | 7 033(23.9) | 18 644(23.0) | 25 677(23.3) |
| Three or four times a week, n(%) | 7 935(27.0) | 21 272(26.3) | 29 207(26.4) |
| Once or twice a week, n(%) | 7 406(25.1) | 20 598(25.4) | 28 004(25.4) |
| One to three times a month, n(%) | 2 986(10.1) | 8 746(10.8) | 11 732(10.6) |
| Special occasions only or never, n(%) | 2 544(8.7) | 7 208(8.9) | 9 752(8.8) |
| Never, n(%) | 1 520(5.2) | 4 548(5.6) | 6 068(5.5) |
| **Smoking status** |  |  |  |
| Never smoker, n(%) | 16 289(55.4) | 46 743(57.7) | 63 032(57.1) |
| Previous smoker, n(%) | 11 311(38.4) | 27 507(33.9) | 38 818(35.2) |
| Current smoker, n(%) | 1 824(6.2) | 6 766(8.4) | 8 590(7.7) |
| **Physical activity, mean(SD), MET minutes/week** | 2 732(2 561) | 2 443(2 497) | 2 520(2 517) |
| **Fresh fruit, mean(SD), pieces/day** | 2.28(2.3) | 1.80(2.7) | 1.93(2.6) |
| **Raw vegetable, mean(SD), tablespoons/day** | 1.89(3.2) | 1.63(3.3) | 1.70(3.3) |
| **Oily fish consumption (times/week)** |  |  |  |
| <2, n(%) | 23 090(78.5) | 68 596(84.7) | 91 686(83.0) |
| ≥2, n(%) | 6 334(21.5) | 12420(15.3) | 18 754(17.0) |
| **Non-oily consumption fish (times/week)** |  |  |  |
| <2, n(%) | 24 473(83.2) | 68 950(85.1) | 93 423(84.6) |
| ≥2, n(%) | 4 951(16.8) | 12 066(14.9) | 17 017(15.4) |
| **Vitamin supplementation, n(%)** | 11 031(37.5) | 8 025(9.9) | 19 055(17.3) |
| **Mineral and other dietary supplementation, n(%)** | 5 686(19.3) | 5 110(6.3) | 10 796(9.8) |
| **Comorbidities** |  |  |  |
| Hypertension, n(%) | 7 162(24.3) | 17 336(21.4) | 24 498(22.2) |
| Type 2 diabetes, n(%) | 137(0.5) | 370(0.5) | 507(0.5) |
| Renal failure, n(%) | 34(0.1) | 108(0.1) | 142(0.1) |
| Myocardial infarction, n(%) | 539(1.8) | 1 144(1.4) | 1 683(1.5) |
| Stroke, n(%) | 235(0.8) | 626(0.8) | 861(0.9) |
| COPD, n(%) | 54(0.2) | 145(0.2) | 199(0.2) |
| Asthma, n(%) | 3 148(10.7) | 9 107(11.2) | 12 255(11.1) |
| Longstanding illness, n(%) | 8 650(29.4) | 21 932(27.1) | 30 582(27.7) |

\*£18 000=€21 489; \$23 253.
BMI: body mass index; MET: metabolic equivalent of task; COPD: chronic obstructive pulmonary disease; SD: standard deviation.

### 3.2 Fish oil use and outcomes

Compared with non-users, participants with habitual use of fish oil supplements were identified to have the reduced risks of being hospitalized or dying from COVID-19. In the analyses, adjusted for age and sex(model 2), we found associations of fish oil use with a 25% reduced risk of COVID-19 hospital admission ([HR 0.95, 95% CI 0.69-0.83 P<0.001; **Table 2**) and a 31% reduced risk of COVID-19 mortality (HR 0.69 95% CI (0.58-0.83 P<0.001; **Table 2**).In the fully adjusted models(model 3),the adjusted hazard ratios associated with fish oil use were 0.79 (95% CI 0.71-0.88 P<0.001; **Table 2**) for COVID-19 hospital admission; 0.72 (95% CI 0.60-0.87 P<0.001; **Table 2**) for COVID-19 mortality. As for SARS-CoV-2 infection, we divided participants into two groups based on median times to follow-up (median 12.1 years). For the group with less than 12.1 years of follow-up, the associations with fish oil use were 0.95(95% CI 0.93-0.98 P=0.002; **Table 2**) in age-and-sex-adjusted model and 0.96(95% CI 0.93-0.99 P=0.019; **Table 2**) in fully adjusted model. No significant associations were observed among participants with more than 12.1 years follow-up. And weighted Kaplan-Meier curves were plotted in **Figure2** for the SARS-Cov-2 infection and COVID-19-related outcomes and showed the fish oil use was associated with reduced risks of COVID-19-related outcomes.

**Table 2.** Associations of use of fish oil with the risk of SARS-CoV-2 infection and COVID-19-related outcomes.

|  | Case/person-<br>years | Non-adjusted model |  | Age and sex-adjusted model |  | Fully adjusted model* |  |
| --- | --- | --- | --- | --- | --- | --- | --- |
|  |  | HR (95% CI) | P | HR (95% CI) | P | HR (95% CI) | P |
| SARS-CoV-2 infection |  |  |  |  |  |  |  |
| Fish oil non-user | 33 935/412007 | 1.00 (Reference) |  | 1.00 (Reference) |  | 1.00 (Reference) |  |
| Fish oil user |  |  |  |  |  |  |  |
| Time<sup>#</sup><12.1 years | 6 111/69 718 | 1.03(1.00-1.06) | 0.033 | 0.95(0.93-0.98) | 0.002 | 0.96(0.93-0.99) | 0.019 |
| Time≥12.1 years | 6 851/87 844 | 1.09(1.06-1.12) | <0.001 | 0.99(0.97-1.03) | 0.853 | 1.02(0.99-1.05) | 0.271 |
| COVID-19 hospital admission |  |  |  |  |  |  |  |
| Fish oil non-user | 1 743/20 422 | 1.00 (Reference) |  | 1.00 (Reference) |  | 1.00 (Reference) |  |
| Fish oil user | 590/6 904 | 0.92(0.85-1.01) | 0.122 | 0.75(0.69-0.83) | <0.001 | 0.79(0.71-0.88) | <0.001 |
| COVID-19 mortality |  |  |  |  |  |  |  |
| Fish oil non-user | 473/5 573 | 1.00 (Reference) |  | 1.00 (Reference) |  | 1.00 (Reference) |  |
| Fish oil user | 172/2 030 | 0.99(0.84-1.19) | 0.958 | 0.69(0.58-0.83) | <0.001 | 0.72(0.60-0.87) | <0.001 |

\*Adjusted for age, sex, ethnicity (white, other), average total annual household income (<£18 000, £18 000-£30 999, £31 000-£51 999, £52 000-£100 000, >£100 000, and missing), Deprivation Index, body mass index, alcohol consumption(daily or almost daily, three or four times a week, once or twice a week, one to three times a month, special occasions only or never, never), smoking status(never, previous, current), physical activity, fresh fruit consumption, raw vegetable consumption, oily fish consumption (<2 or ≥2 times/week), non-oily fish consumption (<2 or ≥2 times/week), vitamin supplement use (yes or no), mineral and other dietary supplement use (yes or no), hypertension (yes or no), type 2 diabetes(yes or no), renal failure(yes or no), myocardial infarction (yes or no), stroke (yes or no), chronic obstructive pulmonary disease (yes or no), asthma (yes or no), longstanding illness (yes or no)
<sup>#</sup>follow-up time
HR: hazard ratio; CI: confidence interval; COVID-19: coronavirus disease 2019

**Figure 2.**
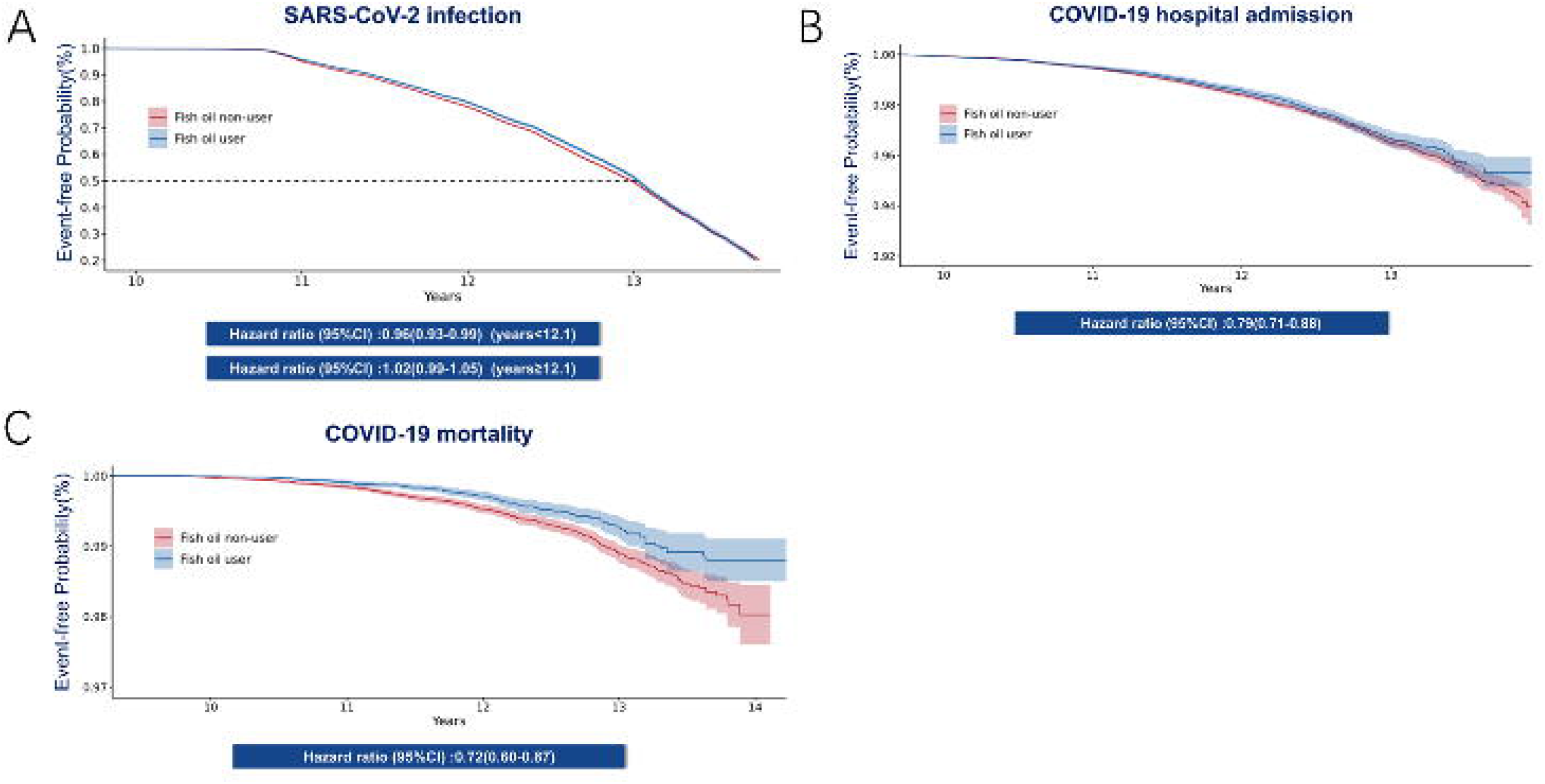
Weighted Kaplan-Meier Curve for fish oil users and non-users in SARS-CoV-2 infection, and COVID-19-related outcomes. The event-free probabilities and hazard ratios for SARS-CoV-2 infection **(A)**, hospital admission **(B)** and death **(C)** with COVID-19 in fish oil users as compared with fish oil non-users are illustrated. 95% confidence intervals are shown in parentheses.

### 3.3 Propensity score-matching analysis

After the match of 29 424 fish oil users and 29 424 non-users, baseline characteristics were more similar (**supplementary Table S1**), except vitamin supplementation (SMD 0.233) and mineral and other dietary supplementation (SMD 0.138). Those who regularly used fish oil supplements were observed with reduced risks for COVID-19 hospital admission (fully adjusted HR 0.77,95% CI 0.69-0.86, P<0,001) and COVID-19 mortality (fully adjusted HR 0.70, 95% CI 0.57-0.85, P<0.001). For SARS-CoV-2 infection, the fish oil users with less than 12.1 years follow-up times were also observed with reduced risks (fully adjusted HR 0.96,95% CI 0.93-0.99, P=0.033) and no associations were identified for those with more than 12.1 years times, which were basically consistent with the results from Cox hazard proportional regression models(**supplementary Table S2**).

### 3.3 Subgroup analysis and sensitivity analyses

Stratified analyses were conducted according to potential risk factors (**Figure 3**). For COVID-19-related outcomes, the association between use of fish oil and the risk of COVID-19 hospital admission were stronger among participants with regular mineral and other dietary supplement (P for interaction =0.036) and who self-reported with myocardial infraction (P for interaction =0.034) or longstanding illness (P for interaction = 0.046). For COVID-19 mortality, the association were stronger among participants with more fresh fruit intake (P for interaction = 0.036) and longstanding illness (P for interaction = 0.0167). No other significant interactions were observed (all P for interaction≥0.05) Logistic regression was used to perform sensitivity analyses. In adjusted models for age and sex, the OR for SARS-CoV-2 infection was 0.97(95% CI 0.93-0.99, P=0.013), for COVID-19 hospital admission was 0.77(0.70-0.85, P <0.001) and for COVID-19 mortality was 0.71(0.59-0.85, P<0.001). In fully adjusted models, the OR for COVID-19 hospital admission was 0.81(0.73-0.90, P <0.001) and for COVID-19 mortality was 0.75(0.62-0.90, P=0.003) But no significant association was observed between SARS-CoV-2 infection and fish oil use (**supplementary Table S3**). In general, these results showed no significant difference in Cox regression models.

**Figure 3.**
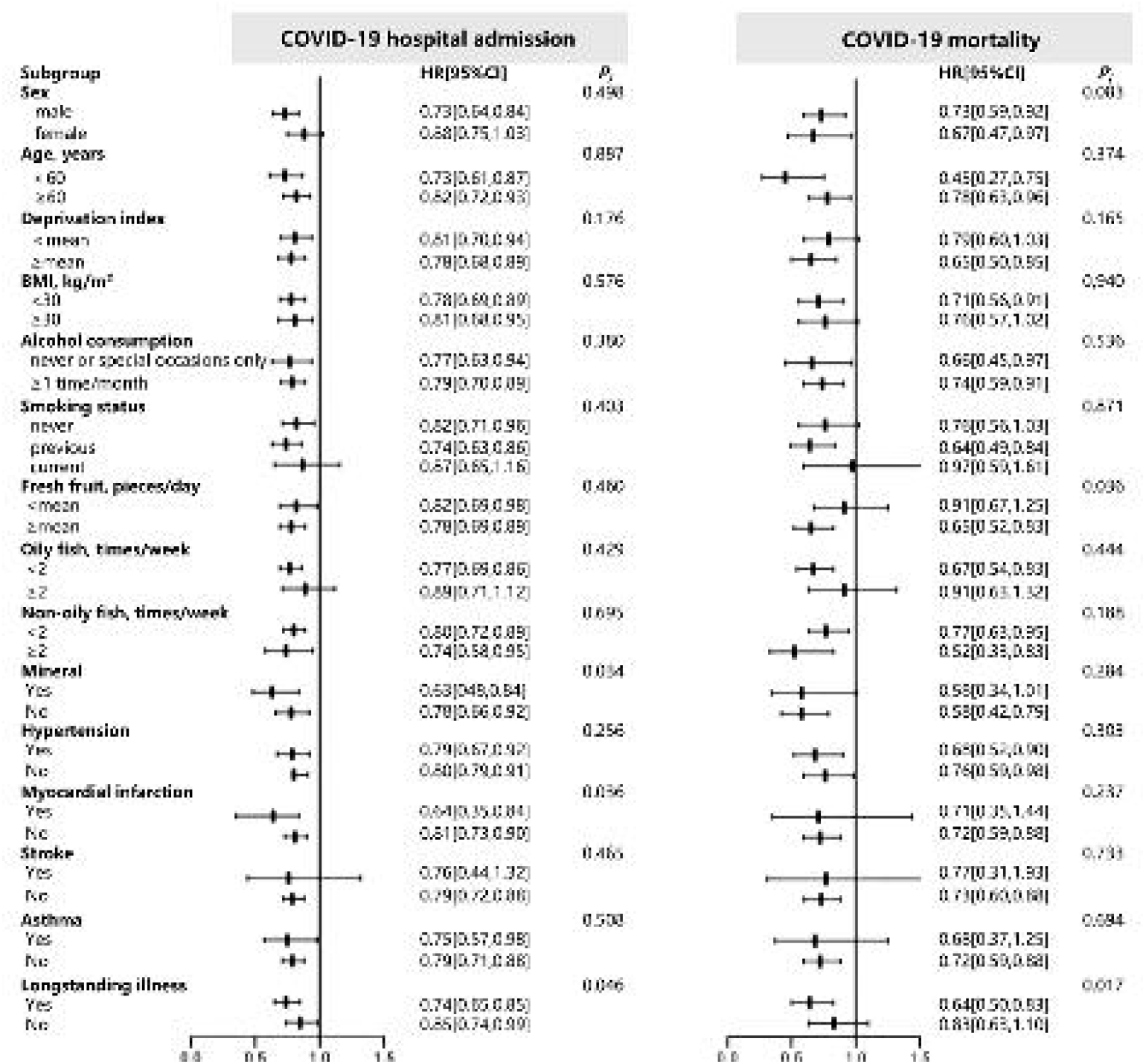
Stratified analysis of fish oil regular users and risk of COVID-19-related outcomes. Effect estimates were based on sex, age, Deprivation Index, body mass index, smoking status, alcohol consumption, smoking status, fresh fruit consumption, oily fish consumption, non-oily fish consumption, mineral and other dietary supplement use, hypertension, myocardial infarction, stroke, asthma, longstanding illness. Cl: confidence interval; HR: hazard ratio; P: P value for interaction.

## 4. Discussion

In this large-scale prospective cohort study, we found that habitual fish oil use was consistently associated with a lower risk of hospital admission and mortality with COVID-19, using fully-adjusted Cox proportional hazard models or logistic regression models. Additionally, no association was found between habitual fish oil use and the risk of COVID-19 infection in the population with more than 12.1 years of follow-up.

### 4.1 Comparison with other studies

To the best of our knowledge, our study is the first large-scale prospective cohort study, utilizing the latest data from the UK biobank, to examine the effects of habitual fish oil use on the risks of SARS-CoV-2 infection, hospital admission and mortality with COVID-19. Our findings are in agreement with the results of several published studies, while their inadequate sample sizes and poor study designs might have limited the power to detect the effects of fish oil, that it plays a protective role in hospital admission and mortality with COVID-19^17 18^.

However, we found that there was no association between habitual fish oil use and the risk of SARS-CoV-2 infection, which is different from previous studies. In Louca’ s study, regular fish oil user had a 12% lower risk of SARS-CoV-2 infection in the UK^19^. One possible explanation is that it was conducted at a time when the omicron variant did not appear. By Jan 10, 2022, omicron cases represented more than 99% of all sequenced cases in the UK^20^. Thus, our analysis of SARS-CoV-2 infection, utilizing the latest COVID-19 data (Version: Feb 2022), found different influences of fish oil use on SARS-CoV-2 infection in different periods. In the earlier stage, fish oil performed a protective function in preventing SARS-CoV-2 infection, which is in accordance with the finding of Louca’ s study. In contrast, there is no association between habitual fish oil use and SARS-CoV-2 infection in the later stage when the Omicron is the dominant variant causing the pandemic in the UK. With high number of mutations, the omicron has novel biological and epidemiological characteristics of immune evasion and high transmissibility, which may account for the failure of protective effect on SARS-CoV-2 infection derived from fish oil use^21^. Since available prevention approaches, such as vaccines, are less effective against the Omicron variant, early preventive steps should be developed to suppress the Omicron variant^22^.

As new variants of SARS-CoV-2 will likely continue to emerge, further large-scale randomized controlled trials of habitual fish oil use testing its effects on COVID-19-related outcomes in existing variants are warranted.

### 4.2 Biological plausibility

There are certain potential mechanisms for the observed benefits for COVID-19-related outcomes derived from fish oil. SARS-CoV-2 infection can cause exaggerated host immune responses, resulting in cytokine storm, the uncontrolled release of proinflammatory cytokines^23^. n-3 PUFAs, the main component of fish oil, have been shown to have powerful anti-inflammatory properties and play a critical role in regulating inflammatory processes through several diverse mechanisms, including acting as substrates for the synthesis of protectins during viral and bacterial infections^24-26^. Moreover, accumulative studies demonstrated that n-3 PUFAs help to attenuate the uncontrolled immune response in the lungs secondary to bacterial or viral infections which could be helpful in the setting of COVID-19^27^. It is also suggested that the immune modulatory properties of n-3 PUFAs will provide significant effects in improving clinical outcomes of COVID-19, particularly in hospitalized high-risk populations with hypertensive, oncologic, and diabetic patients^28^.

In the stratified analysis, we found that the protective associations of fish oil use against hospital admission and mortality with COVID-19 were stronger in those with longstanding diseases. The evidence mentioned below could account for the finding. Several trials found that people with severe underlying conditions, including those pre-existing condition, have disturbed inflammatory components when suffering COVID-19, which may be greatly attenuated by n-3 PUFA^29 30^.

### 4.3 Strengths and limitations of this study

Our study has several strengths. Firstly, a major strength was that we used various methods in statistical analyses, including Cox proportional hazards regression, propensity score-matching analyses, and multivariable logistics regression, to analyze the large-scale cohort with 110,440 participants involved, which demonstrates the effectiveness and adequate statistical power of fish oil use in a real-life setting. Secondly, continuously updated data from the UK biobank allows us to analyze the role of fish oil in the pandemic caused by new variants. Besides, the records of COVID-19 outcomes of UK biobank were directly taken from test results, inpatient hospital data, and death registers to reduce selection bias^14^. Thirdly, detailed information on baseline characteristics and other covariates was available. Thus, we are able to minimize comprehensive confounding factors through constructing fully-adjusted models during the analyses and performing sensitivity analyses.

Several limitations should also be noted. First, the lack of information on the specific dose and frequency of fish oil use from the UK biobank prevented us from assessing potential causal relationship. Moreover, participants of the UK Biobank could not perfectly represent the UK population while they are more representative of healthier population than average, since the individuals were recruited on the premise of voluntariness^31^.

## 5. Conclusions

In conclusion, this large-scale population-based prospective cohort study supported a potentially beneficial association between habitual fish oil use and the risk of hospital admission and mortality with COVID-19, whereas no decreased risk is observed for SARS-CoV-2 infection in the population with more than 12.1 years of follow-up. Further large-scale randomized controlled studies are warranted to confirm the effect of fish oil.

## Data Availability

All data produced in the present work are contained in the manuscript.

## Acknowledgement

The authors thank the UK Biobank for the access of data, and this research has been performed under approval (Application Number 83339).

## Author Contributions

YYM, LJZ, RJZ, and DLL contributed equally to this work. HC, WHS, JHW, and FWL are senior and corresponding authors who also contributed equally to this study. RJZ, WHS, and HC contributed to data extraction, data analyses, and manuscript drafting. YYM, LJZ, and DLL contributed to data interpretation and manuscript drafting. RJ, HHW, ZWZ, QY, and JWL contributed to manuscript drafting. HC, WHS, JHW, and FWL contributed to study design, data interpretation, and final approval of the manuscript. The corresponding author attests that all listed authors meet authorship criteria and that no others meeting the criteria have been omitted.

