## Supplementary Table for "Associations of habitual fish oil use with risk of SARS-CoV-2 infection and COVID-19-related outcomes in UK: national population based cohort study"

**Supplementary Table S1. The demographic and clinical characteristics of patients after propensity score-matching**

| Characteristics | Fish oil user | Fish oil non-user | Overall | SMD |
|---|---|---|---|---|
| **Number of participants, n(%)** | 29 424 | 29 424 | 58 848 | |
| **Age, mean(SD), years** | 58.1 (7.0) | 58.4 (7.4) | 58.2 (7.2) | 0.035 |
| **Sex, female, n(%)** | 16 589 (56.4) | 16 109 (54.7) | 32 698 (55.6) | 0.033 |
| **Ethnicity, White, n(%)** | 28 710 (97.6) | 28 751 (97.7) | 57 461 (97.6) | 0.009 |
| **Household income (£)** | | | | 0.072 |
| <18 000, n(%) | 4 389 (14.9) | 4 553(15.5) | 8 942(15.2) | |
| 18 000-30 999, n(%) | 7 061 (24.0) | 6 659 (22.6) | 13 720 (23.3) | |
| 31 000-51 999, n(%) | 7 479 (25.4) | 7 107 (24.2) | 14 586 (24.8) | |
| 52 000-100 000, n(%) | 5 902 (20.1) | 5 796 (19.7) | 11 698 (19.9) | |
| >100 000, n(%) | 1 478 (5.0) | 1 695 (5.8) | 3 173 (5.4) | |
| Missing, n(%) | 3 115 (10.6) | 3 614 (12.3) | 6 729 (11.4) | |
| **Deprivation index, mean(SD)** | -1.93 (2.7) | -1.96 (2.7) | -1.94 (2.7) | 0.013 |
| **BMI, mean(SD), kg/m$^2$** | 26.58 (4.3) | 26.58 (4.3) | 26.58 (4.3) | 0.002 |
| **Alcohol consumption** | | | | 0.049 |
| Daily or almost daily, n(%) | 7 033 (23.9) | 7 501 (25.5) | 14 534 (24.7) | |
| Three or four times a week, n(%) | 7 935 (27.0) | 7 716 (26.2) | 15 651 (26.6) | |
| Once or twice a week, n(%) | 7 406 (25.2) | 7 171 (24.4) | 14 577 (24.8) | |
| One to three times a month, n(%) | 2 986 (10.1) | 2 814 (9.6) | 5 800 (9.9) | |
| Special occasions only or never, n(%) | 2 544 (8.6) | 2 543 (8.6) | 5 087 (8.6) | |
| Never, n(%) | 1 520 (5.2) | 1 679 (5.7) | 3 199 (5.4) | |
| **Smoking status** | | | | 0.036 |
| Never smoker, n(%) | 16 289 (55.4) | 16 401 (55.7) | 32 690 (55.5) | |
| Previous smoker, n(%) | 11 311 (38.4) | 10 974 (37.3) | 22 285 (37.9) | |
| Current smoker, n(%) | 1 824 (6.2) | 2 049 (7.0) | 3 873 (6.6) | |
| **Physical activity, mean(SD), MET minutes/week** | 2 732(2 561) | 2 773(2 750) | 2 753(2 657) | 0.016 |
| **Fresh fruit, mean(SD), pieces/day** | 2.3 (2.3) | 2.3 (2.2) | 2.3 (2.2) | 0.009 |
| **Raw vegetable, mean(SD), tablespoons/day** | 1.89 (3.2) | 1.86 (3.2) | 1.87 (3.2) | 0.011 |
| **Oily fish consumption (times/week)** | | | | 0.004 |
| <2, n(%) | 23 090 (78.5) | 23 144 (78.7) | 46 234 (78.6) | |
| ≥2, n(%) | 6 334 (21.5) | 6 280 (21.3) | 12 614 (21.4) | |
| **Non-oily consumption fish (times/week)** | | | | 0.003 |
| <2, n(%) | 24 473 (83.2) | 24 504(83.3) | 48 977(83.2) | |
| ≥2, n(%) | 4 951 (16.8) | 4 920(16.7) | 9 871 (16.8) | |
| **Vitamin supplementation, n(%)** | 11 031 (37.5) | 7 853(26.7) | 18 884 (32.1) | 0.233 |
| **Mineral and other dietary supplementation, n(%)** | 5 686 (19.3) | 4 177 (14.2) | 9 863 (16.8) | 0.138 |
| **Comorbidities** | | | | |
| Hypertension, n(%) | 7 162 (24.3) | 7 448 (25.3) | 14 610 (24.8) | 0.023 |

| | | | | |
|---|---|---|---|---|
| Type 2 diabetes, n(%) | 137 (0.5) | 138 (0.5) | 275 (0.5) | <0.001 |
| Renal failure, n(%) | 34 (0.1) | 29 (0.1) | 63 (0.1) | 0.005 |
| Myocardial infarction, n(%) | 539 (1.8) | 634 (2.2) | 1 173 (2.0) | 0.023 |
| Stroke, n(%) | 235 (0.8) | 247 (0.8) | 482 (0.8) | 0.005 |
| COPD, n(%) | 54 (0.2) | 56 (0.2) | 110 (0.2) | 0.002 |
| Asthma, n(%) | 3 148 (10.7) | 3 162 (10.7) | 6 310 (10.7) | 0.002 |
| Longstanding illness, n(%) | 8 650 (29.4) | 8 699 (29.6) | 17 349 (29.5) | 0.004 |

SMD: standard mean difference; BMI: body mass index; MET: metabolic equivalent of task; COPD: chronic obstructive pulmonary disease; SD: standard deviation.

**Supplementary Table S2. Propensity score-matched analysis**

| | Case/person-years | Unadjusted model | | Multivariable-adjusted model* | |
|---|---|---|---|---|---|
| | | HR (95%CI) | *P* | HR (95%CI) | *P* |
| **SARS-CoV-2 infection** | | | | | |
| Fish oil non-user | 13 219/160 063 | 1.00(Reference) | | 1.00(Reference) | |
| Fish oil user | | | | | |
| Time[#]<12.1 years | 6 111/69 718 | 0.96(0.93-0.99) | 0.037 | 0.96(0.93-0.99) | **0.033** |
| Time≥12.1 years | 6 851/87 844 | 0.99(0.96-1.02) | 0.640 | 1.00(0.97-1.04) | 0.856 |
| **COVID-19 hospital admission** | | | | | |
| Fish oil non-user | 743/8 687 | 1.00(Reference) | | 1.00(Reference) | |
| Fish oil user | 590/6 904 | 0.76(0.68-0.85) | **<0.001** | 0.77(0.69-0.86) | **<0.001** |
| **COVID-19 mortality** | | | | | |
| Fish oil non-user | 242/2 830 | 1.00(Reference) | | 1.00(Reference) | |
| Fish oil user | 172/2 030 | 0.68(0.56-0.83) | **<0.001** | 0.70(0.57-0.85) | **<0.001** |

*Adjusted for vitamin supplementation, and mineral and other dietary supplementation

[#]follow-up time

HR: hazard ratio; CI: confidence interval; COVID-19: coronavirus disease 2019

**Supplementary Table S3. Odds ratios (95% CI) for the associations of use of fish oil with the risk of SARS-CoV-2 infection and COVID-19-related outcomes**

| | Case/person-years | Non-adjusted model | | Age and sex-adjusted model | | Fully adjusted model* | |
|---|---|---|---|---|---|---|---|
| | | OR (95% CI) | *P* | OR (95% CI) | *P* | OR (95% CI) | *P* |
| **SARS-CoV-2 infection** | | | | | | | |
| Fish oil non-user | 33 935/412 007 | 1.00 (Reference) | | 1.00 (Reference) | | 1.00 (Reference) | |
| Fish oil user | 12 962/157 562 | 1.09(1.06-1.12) | **<0.001** | 0.97(0.93-0.99) | 0.012 | 0.98(0.95-1.01) | 0.237 |
| **COVID-19 hospital admission** | | | | | | | |
| Fish oil non-user | 1 743/20 422 | 1.00 (Reference) | | 1.00 (Reference) | | 1.00 (Reference) | |
| Fish oil user | 590/6 904 | 0.93(0.84-1.02) | 0.137 | 0.77(0.70-0.85) | **<0.001** | 0.81(0.73-0.90) | **<0.001** |
| **COVID-19 mortality** | | | | | | | |
| Fish oil non-user | 473/5 573 | 1.00 (Reference) | | 1.00 (Reference) | | 1.00 (Reference) | |
| Fish oil user | 172/2 030 | 1.00(0.84-1.20) | 0.989 | 0.71(0.59-0.85) | **<0.001** | 0.75(0.62-0.90) | 0.003 |

*Adjusted for age, sex, ethnicity (white, other), average total annual household income (<£18 000, £18 000-£30 999, £31 000-£51 999, £52 000-£100 000, >£100 000, and missing), Deprivation Index, body mass index, alcohol consumption(daily or almost daily, three or four times a week, once or twice a week, one to three times a month, special occasions only or never, never), smoking status(never, previous, current), physical activity, fresh fruit consumption, raw vegetable consumption, oily fish consumption (<2 or ≥2 times/week), non-oily fish consumption (<2 or ≥2 times/week), vitamin supplement use (yes or no), mineral and other dietary supplement use (yes or no), hypertension (yes or no), type 2 diabetes(yes or no), renal failure(yes or no), myocardial infarction (yes or no), stroke (yes or no), chronic obstructive pulmonary disease (yes or no), asthma (yes or no), longstanding illness (yes or no)

OR: Odds ratios; CI: confidence interval; COVID-19: coronavirus disease 2019
